# First reported case of catheter-related bloodstream infection (CRBSI) caused by *Fereydounia khargensis* in an End-stage kidney disease patient

**DOI:** 10.1101/2024.11.20.24317364

**Authors:** Qiuying Zhang, Mingshui Xie, Yang Liu, Dandan Chen, Wenhang Xie, Min Zhang, Lunhuan Zhou, Zhimin Hu

## Abstract

*F. khargensis* is a yeast and was first described in 2014 from environmental samples. *F. khargensis* belongs to the genus Fereydounia that grows as a yeast, was first identified in East Asia. The basidiomycetous yeast *Fereydounia khargensis* is recorded for the first time from living plants and in East Asia. Phylogenetic analysis indicates a relationship with smut fungi belonging to the order *Urocystidales*, where the monotypic *Fereydouniaceae* and the monogeneric *Doassansiopsidaceae* form the two most basal clades. In culture, this fungus produced cylindrical cells that reproduced by polar budding on short stalks. Production of ballistoconidia and blastospores was observed.

## 1 Introduction

End-stage kidney disease (ESKD) is a leading cause of morbidity and mortality worldwide. An estimated 3.8 million people in the world currently rely on some form of dialysis for treatment of ESKD [1]. Among patients on hemodialysis, infection of hemodialysis accesses (arteriovenous fistulae, arteriovenous grafts, and central venous catheters) was the commonest cause of hospitalisation in 51 (32%) of 159 countries [2]. Catheter-related bloodstream infection (CRBSI) is the most serious complication in patients on hemodialysis with prolonged central venous catheter (CVC) dependence [3]. Phylogenetic analysis indicates a relationship with smut fungi belonging to the order *Urocystidales*, where the monotypic *Fereydouniaceae* and the monogeneric Doassansiopsidaceae form the two most basal clades. *Fereydounia khargensis*, a recently recognized, often multidrug-resistant yeast, has become a significant fungal pathogen due to its ability to cause invasive infections and outbreaks in healthcare facilities.

## 2 Case presentation

After being diagnosed with stage 5 Chronic Kidney Disease (CKD) twelve years ago, a 65-70 year old female patient underwent tunneled central venous catheter placement and received long-term postoperative hemodialysis. 10 days ago, the patient had fever during and after dialysis, with a maximum body temperature of 38.5°C, accompanied by chills. Upon admission (9-26, Day 0), the patient exhibited body temperature of 36.7°C, pulse rate of 89/min, respiratory rate of 21 breaths/min, and blood pressure of 148/90mmHg. She had a medical history of renal hypertension, hypertensive heart disease with heart failure and parathyroidectomy. In the laboratory findings and diagnosis of the patient, various parameters were assessed to understand her overall health status. Blood biochemistry results indicated specific levels such as DD dimer at 776.0ng/mL, myoglobin at 230.0ng/mL, pro-B-type natriuretic peptide (proBNP) at 5590.0pg/mL, ferritin at 243.00ng/mL, calcium at 2.80 mmol/L, phosphorus at 1.84 mmol/L, magnesium at 1.07 mmol/L, urea at 14.54 mmol/L, creatinine at 644.5 μmol/L, uric acid at 298 μmol/L and Estimated Glomerular Filtration Rate (eGFR) at 5.36 mL/min/1.73m^2^. Infection marker PCT examination showed a result of 3.050 ng/ml. Further imaging studies identified the left kidney atrophy and calcified. Cardiac examinations revealed aortic widening, left atrial enlargement (44 mm), left ventricular wall thickening, aortic echogenicity with mild regurgitation, mitral echogenicity with mild regurgitation, minimal tricuspid regurgitation, left ventricular hypodiastolic function (EF: 58%, FS: 30%). The patient underwent peripheral and ductal blood cultures on the first day after admission, both aerobic bottles showing positive after 44.40 hours with peripheral blood culture and 15.36 hours with ductal blood culture. Microscopic examination revealed a Gram-positive retraction septate hypha (Figure 1A). On the second day post-admission, the dialysis catheter was removed and cultured, with results reported on the fourth day, also indicating *F. khargensis*. The specimen was inoculated on a Columbia blood agar (BA), a Chocolate agar (CA) and a CHROMagar Candida plate (CCP, CHROMagar Company, France). Cream coloured, dry and slightly wrinkled growth colonies were observed on the BA and CA after 48 hours (Figure 1B and 1C). The initial colony features are more similar to *Trichosporon spp*.. The colonies appeared blue-gray on chromagar (Figure 1D).

**Fig 1.**
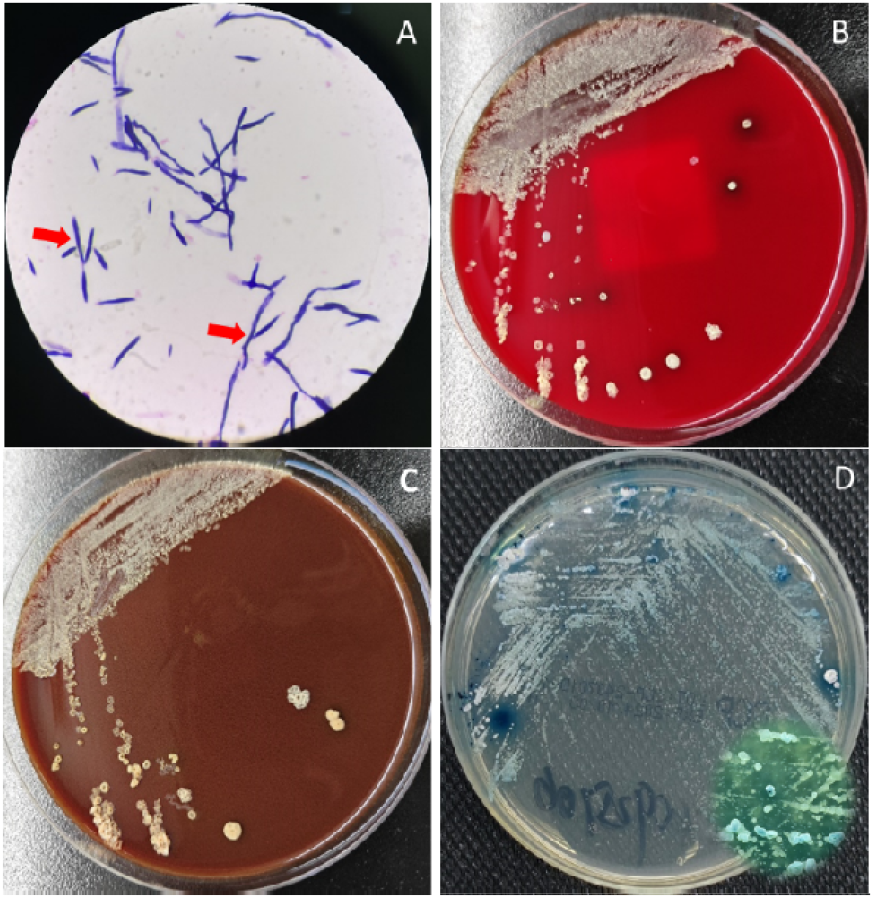
The initial colony characteristics are more similar to Trichosporon spp. cream coloured, dry and slightly wrinkled.

Following diagnosis, the treatment plan was initiate to intravenous injection of 200mg Voriconazole every 12 hours, accompanied by empirical antibacterial treatment to prevent bacterial infections. As the infection symptoms gradually subsided, on the 18th day of hospitalization, the Voriconazole regimen was switched to oral administration of 400 mg daily. Supportive measures, including anti-pressure, metabolic improvement, and anemia correction, collectively contributed to the patient’s improved condition, leading to eventual discharge..

### Mycological Investigations

The yeast-like colonies were subcultured onto new Potato Dextrose Agar (PDA) and incubated at 35□ for further tests. Growths on both PDA plates represented as cream coloured yeast-like colonies, dry and with slightly wrinkled and fringed margins (Fig. 2A). However, after 72 h of incubation, the colonies for both strains started producing melanin-like pigmentation (Fig. 2B) and it became even darker after 120 h of incubation (Fig. 2C). After 48h of incubation on PDA at 35°C, radial powder appeared around the colony, and the center of the colony was in the shape of an umbilical fossa, and the powder around the colony further expanded to the periphery after five days of incubation (Fig. 2D). Slide cultures were performed on PDA and incubated at 35□, fluorescent calcium white staining of 48h isolates revealed short stalk reproduced by polar budding of ellipsoidal and elongation of blastospore (Fig. 3).

**Fig 2.**
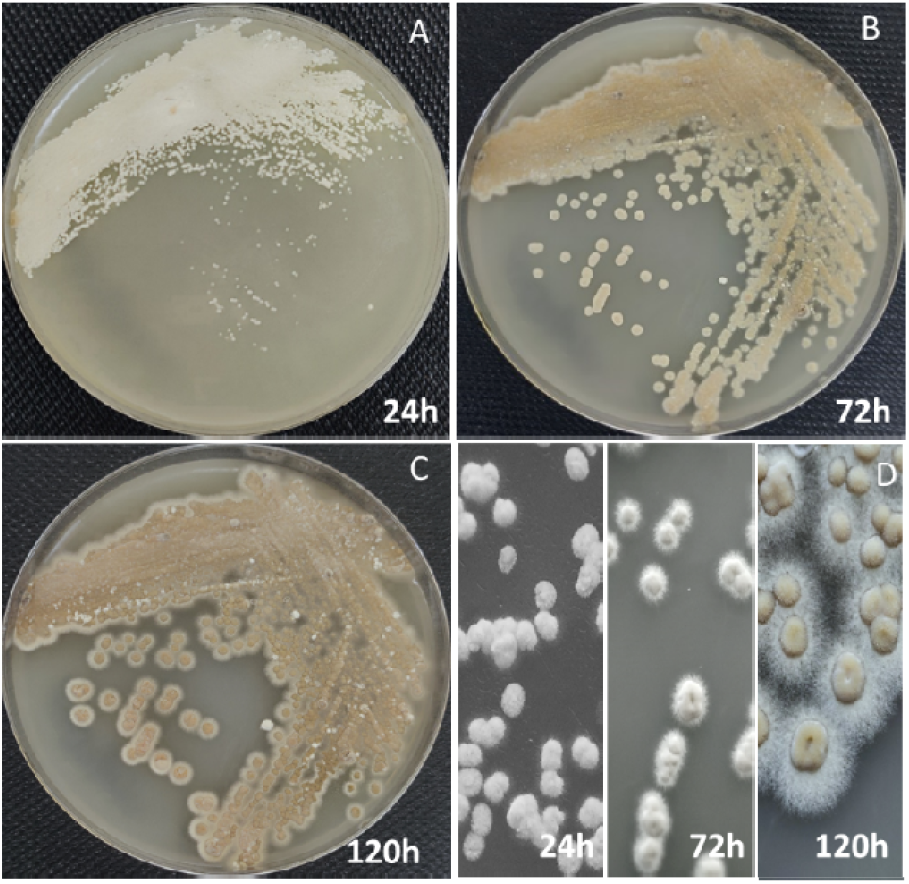
Colonies morphology of yeast-like colony of strain WHY25106 on PDA at 35°C after incubation at 24h (A), 48h (B) and the melanin-like pigment was clearly seen after 120 h incubation (C); Changes in colonies after different times of incubation (D).

**Fig 3.**
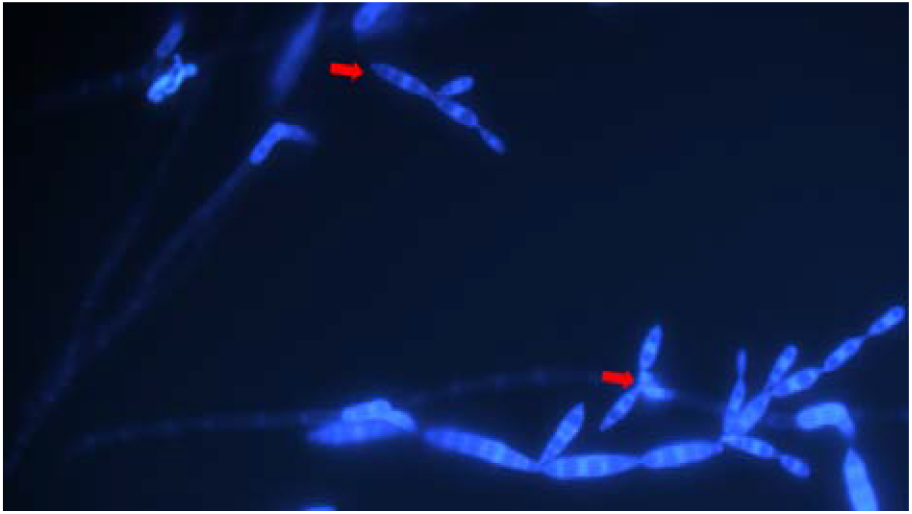
Micromorphology of *F. khargensis* with fluorescent calcium white staining showed vegetative cells without or with blastospores produced by polar budding on short stalks (some budding cells indicated by arrows).

**Fig 4.**
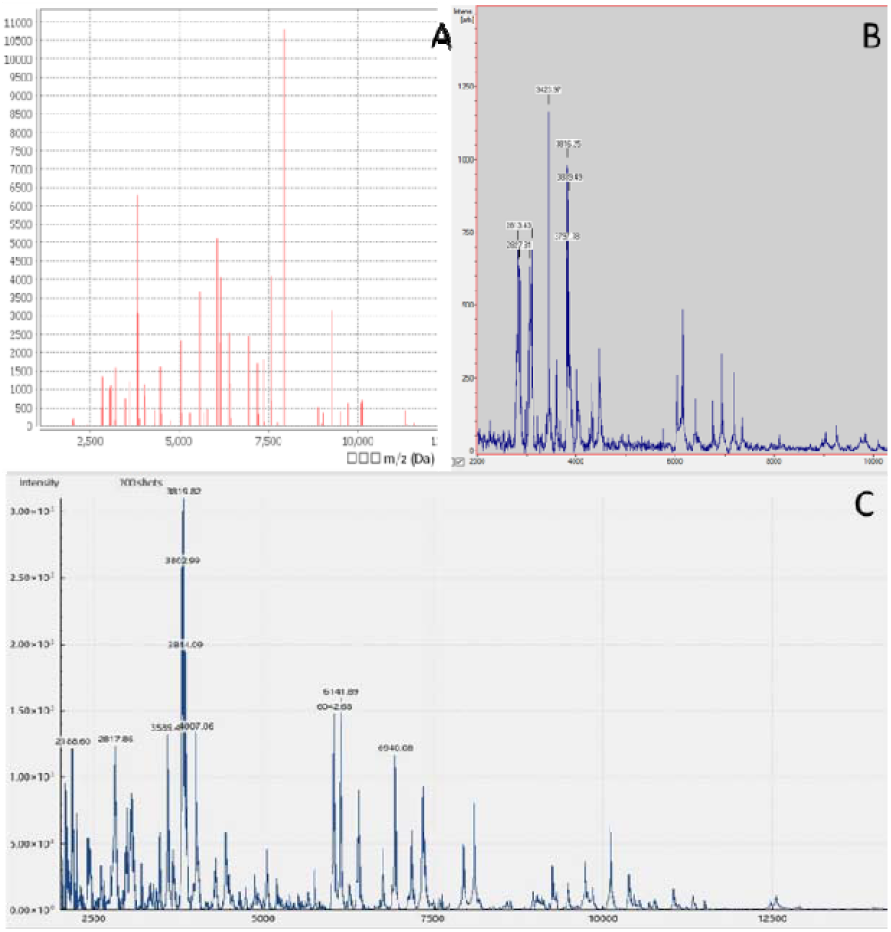
The peak profiles of strain WHY25106 that are generated by the Vitek MS (A), Bruker Biotyper (B), and Zybio EXS2000 (C).

**Fig 5.**
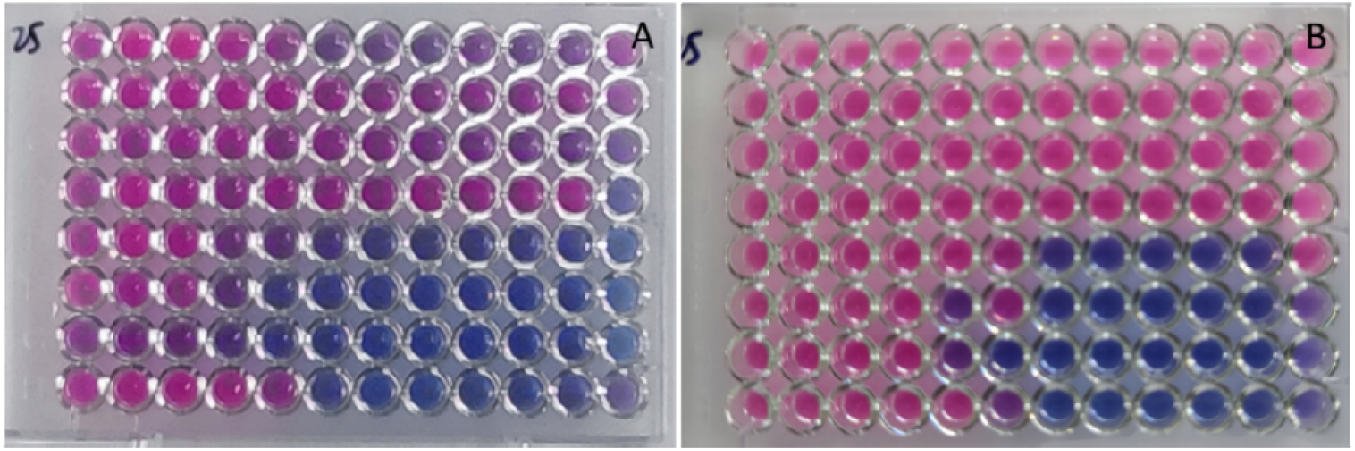
The profiles of SYO10 plates with 24h (A) and 48h (B) incubation

Assimilation tests were carried out by VITEK 2 system (bioMérieux, Marcy-l’étoile) following the manufacturer protocols. The VITEK 2 produced results as *Cryptococcus ciferrii* (50% probability) and *Cryptococcus laurentii* (50% probability), respectively. The following carbon compounds were assimilated: L-malate, erythritol, glycerol, arbutine, D-galactose, gentiobiose, D-glucose, methyl-a-d-glucopyranoside, D-maltose, D-raffinose, D-mannose, D-melezitose, `L-sorbose`, L-rhamnose, xylitol, D-sorbitol, saccharose/sucrose, D-turanose, D-trehalose, nitrate, L-arabinose, D-galacturonate, L-glutamate, D-xylose, DL-lactate, acetate, citrate(sodium), glucuronate, L-proline, 2-keto-d-gluconate, N-acetyl-glucosamine, D-gluconate; no growth occured on amygadline, lactose, D-cellobiose and D-melibiose.

### Identification by MALDI-TOF MS and Molecular Methods

An entire colony was selected and suspended in 300 µL of deionized water containing 900 µL of absolute ethanol. After centrifuging, the pellet was resuspended in 50 µL of 70% formic acid and mixed in a vortex with 50 µL of acetonitrile. Following centrifugation, 1 µL of the clarified lysate was spotted onto the MALDI target plate and air-dried at room temperature. After that, each spot was overlaid with 1 µL of HCCA matrix and completely air-dried before the MALDI-TOF MS measurement. The data analyses were performed using the Bruker BioTyper 3.0 system (Bruker Daltonics), Vitek MS V3.0 system (bioMérieux, Marcy-l’étoile) and Zybio EXQ2000 system (Zhongyuan Huiji Biotechnology). (Fig. 3).

The genomic DNA of isolate was extracted and internal transcribed spacers (ITS) for identifications was amplified using universal primers ITS1/ITS4 as previously described [4]. The nucleotide sequence results were analyzed using BLAST searches (http://blast.ncbi.nlm.nih.gov/); resulting in a 100.0 % matched to the reference strain, IBRC-M30116 (Table 1).

**Table 1.** The reported isolations of *F. khargensis* worldwide.

| Year | Strain name | Sample source | ITS GenBank | Reference |
| --- | --- | --- | --- | --- |
| 2014 | IBRC-M30116 | Plant remnants | KJ490642 | [5] |
| 2016 | UZ1799/14/UZ1801/15 | Human blood/Human pleural | KP326581/KU212883 | [6] |
| 2018 | R. Kirschner 4108/4117/4109 | Lotus leaves | KY400009/KY400010 | [7] |
| 2020 | NR | water | NR | [8] |
| 2021 | NR | The oral washes (non-bedridden nursing home residents) | NR | [9] |
| 2023 | AYC58 | Medical Clinic/Doctor's office | KJ490644 | [10] |
| 2024 | WHY25106 | Human blood | PQ463325/PQ463326 | This report |

NR: Not report.

### In vitro antifungal susceptibility testing (AFST)

AFST was carried out using the custom formulated Sensititre YeastOne One (SYO) microdilution plate (ThermoFisher Scientific, USA). Inoculum suspensions of isolate was prepared according to manufacturer’s protocols. Adjusted suspensions were diluted in YeastOne inoculum broth (ThermoFisher Scientific) and 100 mL dispensed into each well of the SYO10 and incubated for 24 and 48 h, and the results are shown in Table 2. The MIC endpoint was defined as the lowest concentration that produced a color change from pink to blue (amphotericin B) or purple (all other drugs). In vitro susceptibility pattern showed that itraconazole, voriconazole and posaconazole have good activity against the isolate.

**Table 2.** MIC values of F. khargensis against different antifungal agents (μg/mL)

| Antifungal agent | MIC ($\mu\text{g/mL}$) | |
| --- | --- | --- |
|  | 24h | 48h |
| Amphotericin B | 0.5 | 4 |
| Anidulafungin | 0.25 | $\geq 8$ |
| Caspofungin | 2 | $\geq 8$ |
| Micafungin | 8 | $\geq 8$ |
| Flucytosine | 64 | $\geq 64$ |
| Fluconazole | 4 | 4 |
| Itraconazole | 0.03 | 0.25 |
| Voriconazole | 0.06 | 0.5 |
| Posaconazole | 0.06 | 0.5 |

## Disscussion

Invasive fungal infections are a growing threat to immunocompromised patients, highlighting the importance of monitoring fungal pathogens. Emerging pathogens are, in some cases, resistant to the available antifungals, potentiating the threat of novel fungal diseases. Over the last decades, the world has experienced an increase in the incidence and spread of emerging fungal infections. These events may be associated with several factors, including the increase in life expectancy, which generates older individuals with a weaker immune response, as well as increased epigenetic abnormalities that may favor disease establishment. In the case of fungal infections, the reduction in the host’s natural defenses (caused by disease or immunosuppressive medication) meant that numerous pathogens previously considered harmless gradually became disease agents, causing serious infections that are both resistant to antifungal and fatal. Enviromental fungi that are pathogenic to humans exist in a broad range of geographic areas, but they are more common in warmer regions of the planet, probably due to temperature and humidity restrictions on their growth or propagation [11].

*F. khargensis*, commonly known as smut yeast, belongs to the Ascomycota group, exhibiting diverse morphological characteristics; the common feature is sexual reproductive formation of ascospores. The morphology of colonies and cells agree with that described from unidentified plant remains in Iran. According to our observation, the growth rate of this isolate was slower than most yeasts, and the growth control well was observed the inefficient growth of the isolate. So, as to *F. khargensis*, we deduced that the endpoint of in vitro antifungal susceptibility test should be 48 hours. Nevertheless, the occurrence of opportunistic pathogenic yeasts in surface water indicates a potential public health risk to water users, especially in developing countries where many of the population are immunocompromised due to age, disease or economic disadvantage [8].

## Data Availability

All data produced in the present study are available upon reasonable request to the authors

## Ethics approval and consent to participate

Approval was obtained from the ethics committee of Suizhou Central Hospital (KY-2024-003-01). The procedures used in this study adhere to the tenets of the Declaration of Helsinki.

### Consent for publication

The patient consent for publication all the data expose including images.

